# Increased burden of cardiovascular disease in people with liver disease: unequal geographical variations, risk factors and excess years of life lost

**DOI:** 10.1101/2021.10.07.21264705

**Authors:** Wai Hoong Chang, Stefanie H. Mueller, Sheng-Chia Chung, Graham R. Foster, Alvina G. Lai

## Abstract

**Background:** People with liver disease are at increased risk of developing cardiovascular disease (CVD), however, there has yet been an investigation of incidence burden, risk, and premature mortality across a wide range of liver conditions and cardiovascular outcomes.

**Methods:** We employed population-wide electronic health records (EHRs; from 1998-2020) consisting of almost 4 million adults to assess regional variations in disease burden of five liver conditions, alcoholic liver disease (ALD), autoimmune liver disease, chronic hepatitis B infection (HBV), chronic hepatitis C infection (HCV) and NAFLD, in England. We analysed regional differences in incidence rates for 17 manifestations of CVD in people with or without liver disease. The associations between biomarkers and comorbidities and risk of CVD in patients with liver disease were estimated using Cox models. For each liver condition, we estimated excess years of life lost (YLL) attributable to CVD (i.e., difference in YLL between people with or without CVD).

**Results:** The age-standardised incidence rate for any liver disease was 114.5 per 100,000 person years. The highest incidence was observed in NAFLD (85.5), followed by ALD (24.7), HCV (6.0), HBV (4.1) and autoimmune liver disease (3.7). Regionally, the North West and North East regions consistently exhibited high incidence burden. Age-specific incidence rate analyses revealed that the peak incidence for liver disease of non-viral aetiology is reached in individuals aged 50-59 years. Patients with liver disease had a 2-fold higher incidence burden of CVD (2,634.6 per 100,000 persons) compared to individuals without liver disease (1,339.7 per 100,000 persons). When comparing across liver diseases, atrial fibrillation was the most common initial CVD presentation while hypertrophic cardiomyopathy was the least common. We noted strong positive associations between body mass index and current smoking and risk of CVD. Patients who also had diabetes, hypertension, proteinuric kidney disease, chronic kidney disease, diverticular disease and gastro-oesophageal reflex disorders had a higher risk of CVD, as do patients with low albumin, raised C-reactive protein and raised International Normalized Ratio levels. All types of CVD were associated with shorter life expectancies. When evaluating excess YLLs by age of CVD onset and by liver disease type, differences in YLLs, when comparing across CVD types, were more pronounced at younger ages.

**Conclusions:** We developed a public online app (https://lailab.shinyapps.io/cvd_in_liver_disease/) to showcase results interactively. We provide a blueprint that revealed previously underappreciated clinical factors related to the risk of CVD, which differed in the magnitude of effects across liver diseases. We found significant geographical variations in the burden of liver disease and CVD, highlighting the need to devise local solutions. Targeted policies and regional initiatives addressing underserved communities might help improve equity of access to CVD screening and treatment.

## Background

Although cardiovascular disease (CVD) prevention policies have had some success, for the past two decades, they have remained limited in reducing the number of deaths globally with CVD still ranked as the leading cause of death. A large proportion of deaths are attributable to preventable illnesses - most premature deaths from CVD in people younger than 75 are avoidable^1,2^. As major risk factors for CVD, type 2 diabetes and obesity have received the most attention in policy, public awareness and guidelines. However, despite reported associations between liver disease and cardiovascular risk, liver conditions other than non-alcoholic fatty liver disease (NAFLD) have mostly been overlooked.

Liver disease encompasses a spectrum of conditions including viral hepatitis, NAFLD, steatosis to end-stage cirrhosis. Other causes of liver cirrhosis include autoimmune liver disease and alcoholic liver disease (ALD). The heart has been implicated in the progression of aggravating liver disease, with the liver-heart axis more extensively explored in patients with NAFLD^3^. With a global prevalence of 25% in the general population^4^, the number of individuals living with NAFLD far exceeds the number of individuals with diabetes and obesity^5^ combined. NAFLD is characterised by lipid accumulation in the liver and systemic metabolic aberrations, which leads to an increased risk of developing CVD. However, the pathophysiological associations between other liver diseases and CVD remain poorly understood. The World Health Organisation estimated that 250 million individuals are living with chronic hepatitis B caused by infection with the hepatitis B virus (HBV) where prevalence is the highest in African and Western Pacific regions^6^. An estimated 71 million individuals have chronic hepatitis C virus (HCV) infection^7^ and emerging evidence suggests an association between HCV and CVD, where CVD associated with HCV infection is responsible for the loss of 1.5 million disability-adjusted life-years each year^8^. HBV and HCV transmissions continue to rise in low- and middle-income countries and sustained chronic infection may lead to the development of CVD due to chronic inflammation and metabolic derangements^9^. Additionally, liver disease often co-exists with type 2 diabetes^10^ and chronic renal disease^11^, both of which are independent cardiovascular risk factors^12^.

Current guidelines from the American Association for the Study of Liver Diseases recommended the modification of CVD risk factors in these individuals and the screening of CVD during liver transplant evaluation^13^. Furthermore, since CVD is the most common cause of death in patients with NAFLD, the European Association for the Study of Liver recommends mandatory screening for CVD in individuals with NAFLD^14^. As liver disease progresses, liver-specific risk factors (e.g., raised International Normalised Ratio) and prevalent comorbidities that are independently associated with increased CVD risk may come into play and could result in more severe illness in these individuals. Yet, CVD is often underdiagnosed in patients with liver disease and there are no policies on screening patients with chronic liver disease.

Harnessing an emerging data science opportunity from population-based electronic health records (EHRs), we sought to identify population groups, among patients with liver disease, who might be at high risk for CVD. Employing linked EHRs from primary and secondary care on 4 million individuals, our study aims to address the value, or futility, of targeted monitoring of CVD risk in patients with liver disease. We characterised clinical features of patients with any of the five liver conditions, ALD, autoimmune liver disease, HBV, HCV and NAFLD and explored the associations between risk factors and future risk of 17 of the most common initial cardiovascular presentations. A report on the atlas of variation in risk factors and healthcare for liver disease by the Public Health England found that for the past three decades, there have been limited improvements in mortality rates and incidence of liver disease^15^. Deaths from liver disease have increased by fourfold and progress on earlier diagnosis and better treatment have been slow^16,17^. For these reasons, we investigated regional variations in liver disease burden and CVD burden to highlight potential gaps in the provision of services, inequality of access and prevention initiatives, drawing attention to regions where improvements are most needed. Specific objectives of this study were: (i) to estimate the regional variations in incidence rates for liver disease in England, (ii) to estimate regional variations in incidence rates for CVD in patients with and without liver disease, (iii) to estimate the associations of age, clinical biomarkers, pre-existing comorbidities and smoking with risk of initial presentation of CVD and (iv) to estimate excess years of life lost (a marker of pre-mature mortality) based on the age of CVD onset by comparing patients with liver disease who subsequently developed CVD to those who did not.

We provide an open-access online app, with implications for clinical risk assessment and targeted policy on CVD prevention, that shows incidence rates, years of life lost estimations and cause-specific hazard ratios for associations between clinical features and initial presentation of CVD.

## Methods

### Study design and data source

We used linked EHRs from primary and secondary care, which consisted of a population of 3,929,596 adults aged ≥ 30 years during the study period of 1998 to 2020. Individuals were followed up until the occurrence of a primary endpoint, death, date of last data collection for the practice, date of administrative censoring (June 2020) or deregistration from the practice (i.e., loss to follow-up), whichever occurred first. Baseline characteristics were analysed and stratified by liver disease. Information governance approval was obtained from the Medicines Healthcare Regulatory Authority (UK) Independent Scientific Advisory Committee (21_000363) Clinical Practice Research Datalink (CPRD).

### Open electronic health record (EHR) definitions of diseases and covariates

Phenotype definitions of liver disease, CVD, comorbidities and risk factors are available at https://caliberresearch.org/portal and have previously been validated^18–23^. Phenotypes for primary care records were generated using Read clinical terminology (version 2). Phenotypes for secondary care records were generated using ICD-10 terms. BMI and smoking were considered as the nearest record to entry within 1 year prior to entry. We examined nine biomarkers; albumin, alanine aminotransferase, aspartate transaminase, bilirubin, C-reactive protein, gamma-glutamyltransferase, International Normalised Ratio (INR), platelets and triglycerides. The primary outcome was the first record of one of the following 17 cardiovascular presentations in data from either primary or secondary care, which were grouped into five CVD categories: (i) coronary heart disease: stable angina, unstable angina, myocardial infarction, heart failure and coronary heart disease unspecified; (ii) strokes and transient ischaemic attack (TIA): ischaemic stroke, stroke unspecified and TIA; (iii) peripheral vascular disease: peripheral arterial disease, pulmonary embolism, venous thromboembolic disease and abdominal aortic aneurysm; (iv) cardiomyopathy: dilated cardiomyopathy, hypertrophic cardiomyopathy and cardiomyopathy unspecified; (v) arrhythmia: atrial fibrillation and sick sinus syndrome. We examined 14 comorbidities and considered records for comorbidities prior to cohort entry (prevalent comorbidities). The comorbidities were diabetes mellitus, complications of diabetes (i.e., diabetic nephropathy, diabetic retinopathy and neurological complications of diabetes), dyslipidaemia, hypertension, jaundice, proteinuric kidney disease, oesophagitis or oesophageal ulcer, proteinuria, chronic kidney disease and gastrointestinal conditions such as Barrett’s oesophagus, Crohn’s disease, diverticular disease of the intestine, gastro-oesophageal reflux disease and irritable bowel syndrome.

### Estimations of age-standardised and age-specific incidence rates

Age-standardised incidence rates were estimated per 100,000 person-years and based on a five-year study period from 01.01.2015 till 31.12.2019. We analysed geographical variations in incidence rates based on CPRD practice region definitions. We retrieved the 2019 population estimates (overall and by age groups) from the Office for National Statistics^24^ for age-standardisation of incidence rates. Additionally, we estimated incidence rates by age groups (i.e., age-specific incidence rates) to explore differences across the age groups. Patient years were calculated by age group (<30, 30-39, 40-49, 50-59, 60-69, 70-79 and ≥ 80) using the ‘pyears’ function in the survival package in R (version 3.2.10). Confidence intervals for incidence rates were calculated based on the central limit theorem given dichotomous outcome in a single population. For event numbers smaller than five, age-standardised incidence rate was not reported. Using these approaches, we estimated both age-standardised and age-specific incidence rates for liver disease and incidence rates for CVD in patients with and without liver disease.

### Estimations of excess years life lost (YLL)

YLL was estimated using the lillies package^25^, which was validated by other studies^26–28^. YLL was estimated based on CVD onset at ages 30, 40, 50, 60, 70 and 80. We estimated excess YLL based on the specific age of onset of CVD and compared the average of these individuals to the patients without CVD of the same age. We also examined excess YLL by the type of first CVD presentation within age groups.

### Statistical analyses

Cox proportional hazards models were fitted. Hazard ratios (HRs) from the fully adjusted models were reported with 95% confidence intervals (CI). All P values were two-sided. Models were also refitted with age group as a categorical variable to return HRs by age group. Proportional hazards assumption violations were tested for a zero slope in the scaled Schoenfeld residuals. Biomarker and BMI measurements were indicated as the presence or absence of a particular measurement above or below the stated threshold. In the primary analysis, those with missing BMI information were assumed to be non-obese. Those with missing biomarker information were assumed to be within the normal threshold on the assumption that abnormal blood test results were likely to be recorded if present. Additionally, sensitivity analyses were conducted on complete records to demonstrate that the estimates were robust to our assumptions around missing data.

Data were analysed using R (3.6.3) with the following packages: survival, tidyverse, tableone, epitools, rgdal, broom, ggplot2 and ggmap.

## Results

The study cohort included 3,929,596 individuals, of whom 12,845, 2,210, 1,753, 3,112 and 20,928 individuals were newly diagnosed with ALD, autoimmune liver disease, HBV, HCV and NAFLD, respectively (Additional file 4). The median age of diagnosis for each liver disease was ALD (54.8; IQR: 46.6 - 63.1), autoimmune liver disease (63.3; IQR: 53.1 - 72.6), HBV (47.1; IQR: 38.3 - 57.0), HCV (46.50; IQR: 39.3 - 54.9) and NAFLD (56.8; IQR: 47.8 - 66.6) (Additional file 4). Patient characteristics (i.e., prevalent comorbidities, biomarker values at baseline, general practice region and smoking) stratified by liver disease and sex were described in Additional file 4. Comorbidity patterns in patients with liver disease were analysed and presented in the supplementary appendix and Additional file 1.

### Patients with liver disease had a higher burden of CVD

The age-standardised incidence rate for any of the five liver diseases in England was 114.5 per 100,000 person years (95% confidence interval (CI): 112.5-116.6) (Figure 1A, Additional file 5). For specific liver diseases, the age-standardised incidence rates per 100,000 person years were as follow: ALD (24.7, CI: 23.8-25.7), autoimmune liver disease (3.7, CI: 3.3-4.1), HBV (4.1, CI: 3.7-4.5), HCV (6.0, CI: 5.5-6.4) and NAFLD (85.5, CI: 83.7-87.3) (Figure 1A, Additional file 5). We also estimated age-specific incidence rates for liver disease and observed that the rate for NALFD peaked in individuals aged 50 to 59 years (Figure 1B, Additional file 6). Similar trends were observed in other liver conditions of non-viral aetiology where incidence rates were as follow: ALD (age 50-59), autoimmune liver disease (age 60-69) and NAFLD (age 50-59). However, for HBV and HCV, the highest incidence rates were observed in younger people, individuals aged 30-39 and individuals aged 40-49, respectively (Figure 1B, Additional file 6).

**Figure 1.**
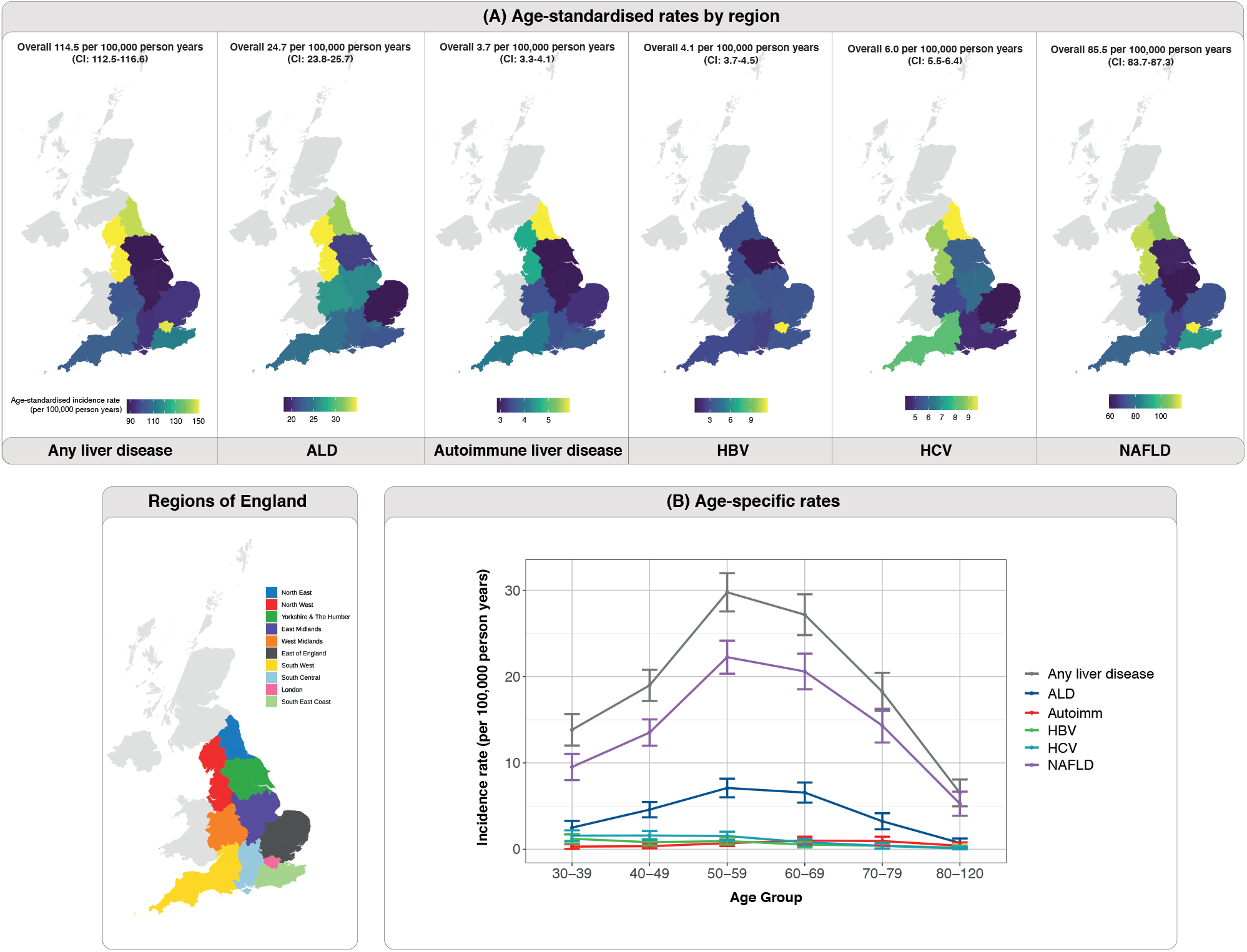
Variations in incidence rates for liver disease. (A) Age-standardised incidence rates by geographical regions. (B) Age-specific incidence rates of liver disease to demonstrate changes in incidence rates across age-groups.

To ascertain whether patients with liver disease had a higher incidence of CVD, we considered 17 cardiovascular conditions (see methods). The age-standardised incidence rate for CVD in patients with any liver disease was two-fold higher than in people without liver disease. Incidence rate was 2,634.6 per 100,000 person years (CI: 2,524.4-2,744.8) in people with liver disease (Figure 2A, Additional file 7) compared to 1,339.7 per 100,000 person years (CI: 1,332.3-1,347.0) in people without liver disease (Figure 2B, Additional file 8). Incidence burden of CVD in patients with specific liver conditions were: ALD (3,173.3; CI: 2,955.3-3,391.3), autoimmune liver disease (2,084.8; CI: 1,731.8-2,437.8), HBV (1,550.0; CI: 1,312.6-1,787.5), HCV (1,999.6; CI: 1,766.4-2,232.7) and NAFLD (2,878.0; CI: 2,708.1-3,048.0) (Figure 2A, Additional file 7).

**Figure 2.**
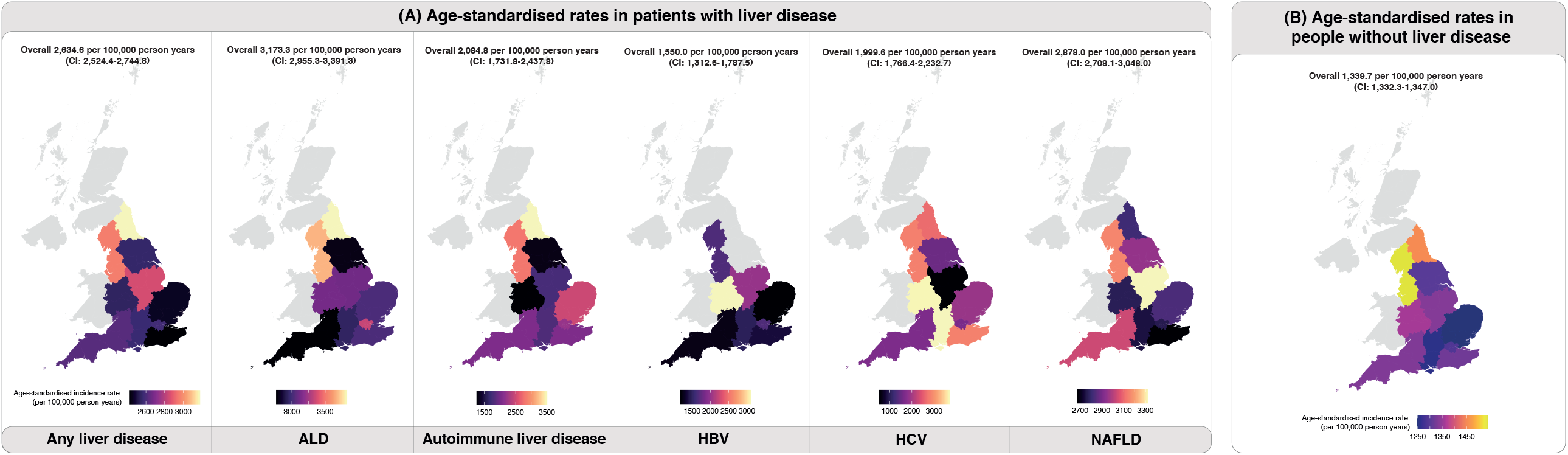
Variations in incidence rates for cardiovascular disease (CVD). Age-standardised incidence rates for CVD by geographical regions in (A) patients with liver disease and (B) patients without liver disease.

The age-specific incidence rates of CVD in patients with any liver disease exhibited an upward trend with increasing age, peaking at ages 70 to 79 (766 per 100,000 person years). Similarly, in people without liver disease, this upward trend was maintained, albeit at a lower magnitude (highest CVD incidence was observed in individuals aged 80 and above; 562 per 100,000 person years) (Additional file 9, Additional file 10). Unlike the incidence rates of CVD which increased with age peaking in the highest age groups, the incidence rates of liver disease peaked in middle-aged individuals as shown earlier (Figure 1B). Geographical variations of liver disease burden and CVD burden (in people with or without liver disease) were further explored and presented in the supplementary appendix.

### Patterns of the first presentation of CVD in patients with liver disease

We analysed the first presentation of 17 types of incident CVD. When considering the first presentation of CVD, the number of incident CVD events for patients with liver disease were as follow; ALD (3,283/12,845, 25.6%), autoimmune liver disease (667/2,210, 30.2%), HBV (270/1,753, 15.4%), HCV (522/3,112, 16.8%) and NAFLD (4,362/20,928, 20.8%). When comparing across patients with different liver diseases, atrial fibrillation was the most common condition while hypertrophic cardiomyopathy was the least common (Figure 3A). When individual CVDs were grouped into five categories, coronary heart disease, which is a composite of stable angina, unstable angina, myocardial infarction, heart failure and coronary heart disease unspecified, was the most common, followed by arrhythmia, stroke/TIA, peripheral vascular disease and cardiomyopathy (Figure 3B). When delving into specific liver conditions, coronary heart disease was the first presentation in 53% of patients with NAFLD who had cardiovascular events, while cardiomyopathy was the first presentation in only 3% of patients. By contrast, 44% of patients with autoimmune liver disease presenting with cardiovascular events had coronary heart disease, and 1% had cardiomyopathy (Figure 3B).

**Figure 3.**
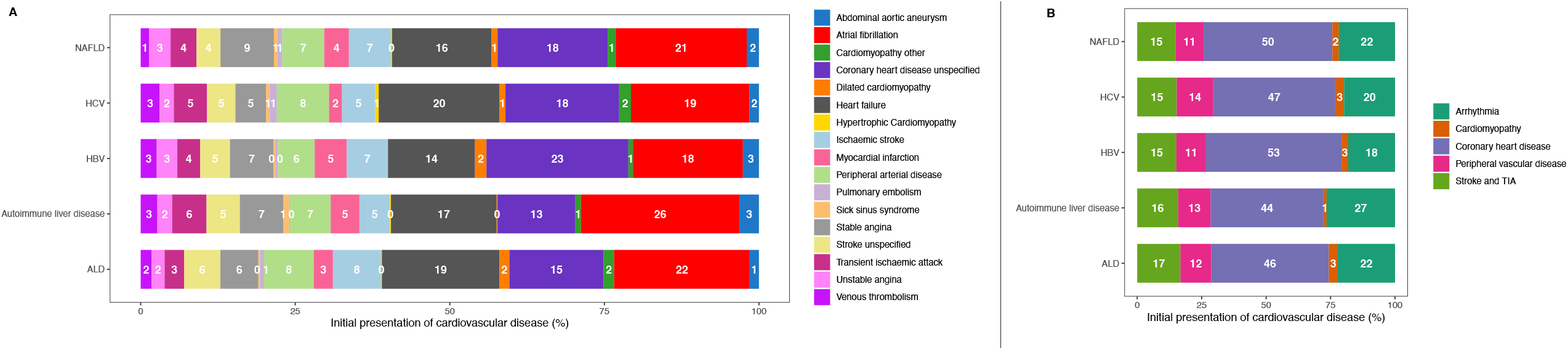
Distribution of initial presentations of incident cardiovascular diseases in patients with liver disease. (A) Distribution of each of the 17 types of cardiovascular diseases. (B) Distribution by five CVD categories.

The first presentation of CVD was influenced by age and sex (Additional file 2). CVD was more common in older individuals and there are sex differences in the type of CVD presented. In patients with HCV, for example, 35% of men and 43% of women aged 70 – 79 had a cardiovascular event (Additional file 2). In men, 5%, 5%, 6% and 19% of these events account for peripheral vascular disease, stroke/TIA, arrhythmia and coronary heart disease respectively. While in women, the proportion differs with 4%, 15% and 24% events accounting for stroke/TIA, arrhythmia and coronary heart disease respectively (Additional file 2).

### Clinical features associated with the first presentation of CVD

The associations between patient-level factors (i.e., prevalent comorbidities, biomarkers and smoking) and the risk of incident CVD were shown in Figure 4 and Additional file 11. We noted strong positive associations between increasing age and risk of incident CVD. Among patients with ALD, individuals aged 60-69 had a 4-fold higher risk compared to 30-39-year-olds (adjusted HR: 4.98, CI: 4.20 – 5.91). The effect of age was consistent across all liver diseases. Women had a lower risk than men; ALD (adjusted HR: 0.72, CI: 0.67 – 0.78), autoimmune liver disease (adjusted HR: 0.53, CI: 0.45 – 0.64), HBV (adjusted HR: 0.67, CI: 0.52 – 0.87), HCV (adjusted HR: 0.73, CI: 0.60 – 0.88) and NAFLD (adjusted HR: 0.71, CI: 0.67 – 0.76) (Figure 4, Additional file 11).

**Figure 4.**
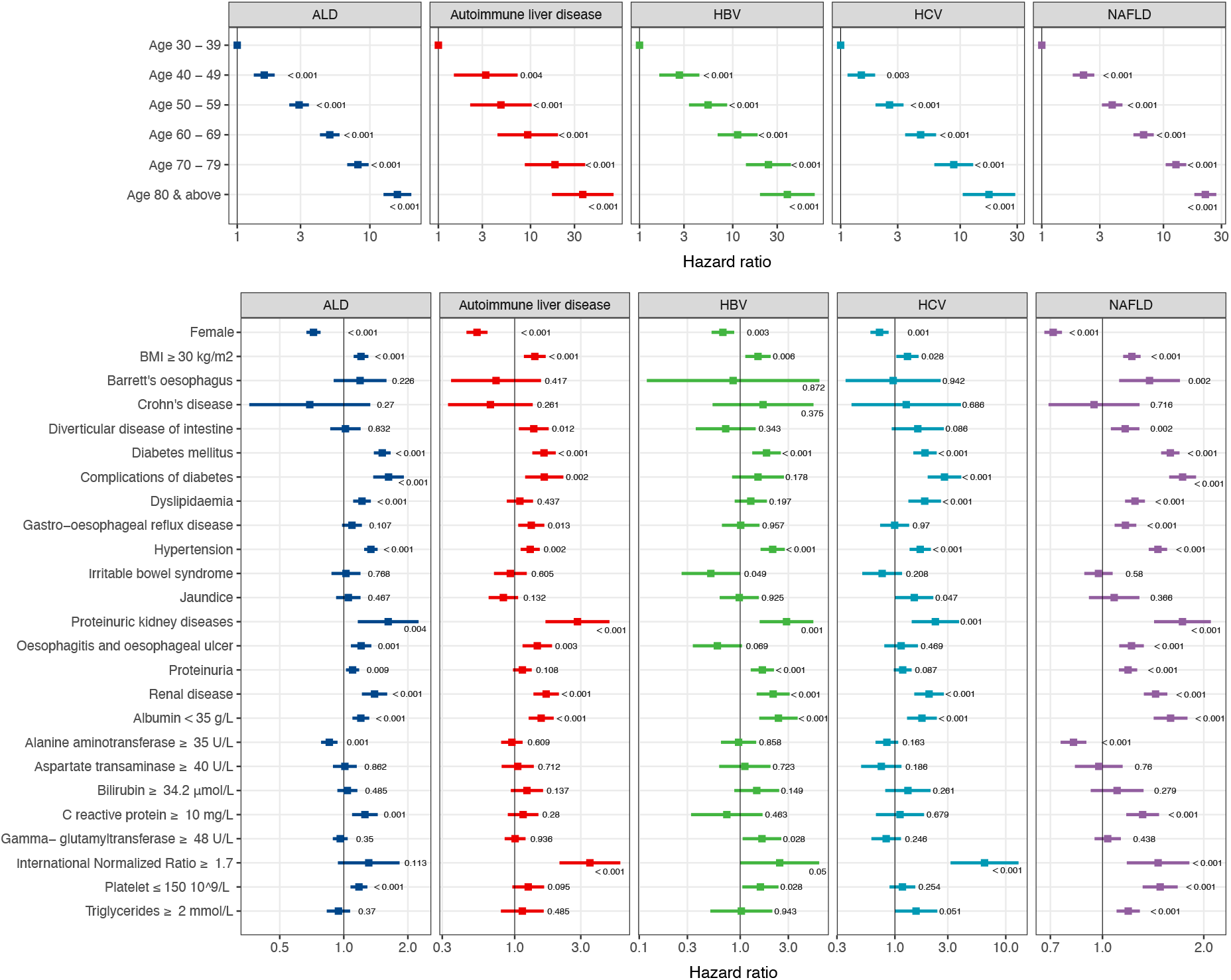
Multivariable Cox model was fitted and fully adjusted for other covariates for the initial presentation of cardiovascular disease in patients with liver disease. Forest plots indicate estimated hazard ratios for each patient characteristic where hazard ratios are plotted on a log scale. Error bars represent confidence intervals for the hazard ratio. P values are indicated on the plot.

We observed a consistent pattern of increased risk for a body mass index (BMI; kg/m^2^) of over 30; ALD (adjusted HR: 1.20, CI: 1.11 – 1.30), autoimmune liver disease (adjusted HR: 1.40, CI: 1.17 – 1.67), HBV (adjusted HR: 1.50, CI: 1.12 – 2.00), HCV (adjusted HR: 1.30, CI: 1.03 – 1.64) and NAFLD (adjusted HR: 1.22, CI: 1.15 – 1.30) (Figure 4, Additional file 11). Current smoking was associated with a higher risk in patients with ALD (adjusted HR: 1.30, CI: 1.18 - 1.43), autoimmune liver disease (adjusted HR: 1.78, CI: 1.45 - 2.18), HBV (adjusted HR: 1.74, CI: 1.25 - 2.43) and NAFLD (adjusted HR: 1.77, CI: 1.64 - 1.91). The same pattern was observed in former smokers in patients with ALD, autoimmune liver disease or NAFLD.

Comorbidities such as diabetes mellitus, complications of diabetes, hypertension, proteinuric kidney disease and chronic renal disease were associated with a higher risk of CVD when comparing across all liver diseases. Diverticular disease and gastro-oesophageal reflex disease were associated with a higher risk of CVD in patients with autoimmune liver disease or NAFLD. Dyslipidaemia was also associated with a higher risk in patients with ALD (adjusted HR: 1.22, CI: 1.11 – 1.34), HCV (adjusted HR: 1.86, CI: 1.32 – 2.60) or NAFLD (adjusted HR: 1.25, CI: 1.17 – 1.34) (Figure 4, Additional file 11).

We observed that albumin levels of < 35 g/L were associated with a higher risk of CVD in patients with ALD (adjusted HR: 1.20, CI: 1.10 - 1.32), autoimmune liver disease (adjusted HR: 1.55, CI: 1.26 - 1.91), HBV (adjusted HR: 2.39, CI: 1.54 – 3.71), HCV (adjusted HR: 1.75, CI: 1.29 – 2.39) or NAFLD (adjusted HR: 1.59, CI: 1.42 - 1.79). In patients with ALD or NAFLD, those who had elevated C-reactive protein levels of ≥ 10 mg/L, a marker of inflammation, had at least a 1.2-fold increased risk of CVD. Raised International Normalized Ratio (INR) of ≥ 1.7 is associated with 3.5-fold, 6.4-fold and 1.5-fold increased risk of CVD in patients with autoimmune liver disease, HCV or NAFLD. Interestingly, we noted an inverse association between alanine aminotransferase (ALT) levels and CVD risk in patients with ALD (adjusted HR: 0.85, CI: 0.78 - 0.93) and NAFLD (adjusted HR: 0.82, CI: 0.75 - 0.90). We performed sensitivity analyses using complete records for BMI, albumin, C-reactive protein and INR and found similar results (Additional file 12). Complete record analysis on ALT levels and CVD risk also revealed inverse associations where patients with raised ALT (≥ 35 U/L) had a lower risk of CVD (reasons for this are explored in the discussion section).

### Excess years of life lost from CVD

Among patients with liver disease, we estimated excess years life lost (YLL), which was calculated as the average number of years that individuals with a specific CVD condition lose in excess of that found in people without CVD of the same sex and age. We estimated excess YLL based on the age of CVD onset at ages 30, 40, 50, 60, 70 and 80 (Figure 5A). Overall, individuals with NAFLD experienced the highest excess YLL upon CVD diagnosis, a pattern that was most pronounced at younger ages. Patients with NAFLD who developed CVD at the age of 40 years experienced an excess YLL of 18.2 years (CI: 16.5 – 20.2) compared to those without CVD. In contrast, at the same age of CVD onset, patients with HBV experienced excess YLL of only 9.5 years (CI: 7.1 – 12.1) (Figure 5A).

**Figure 5.**
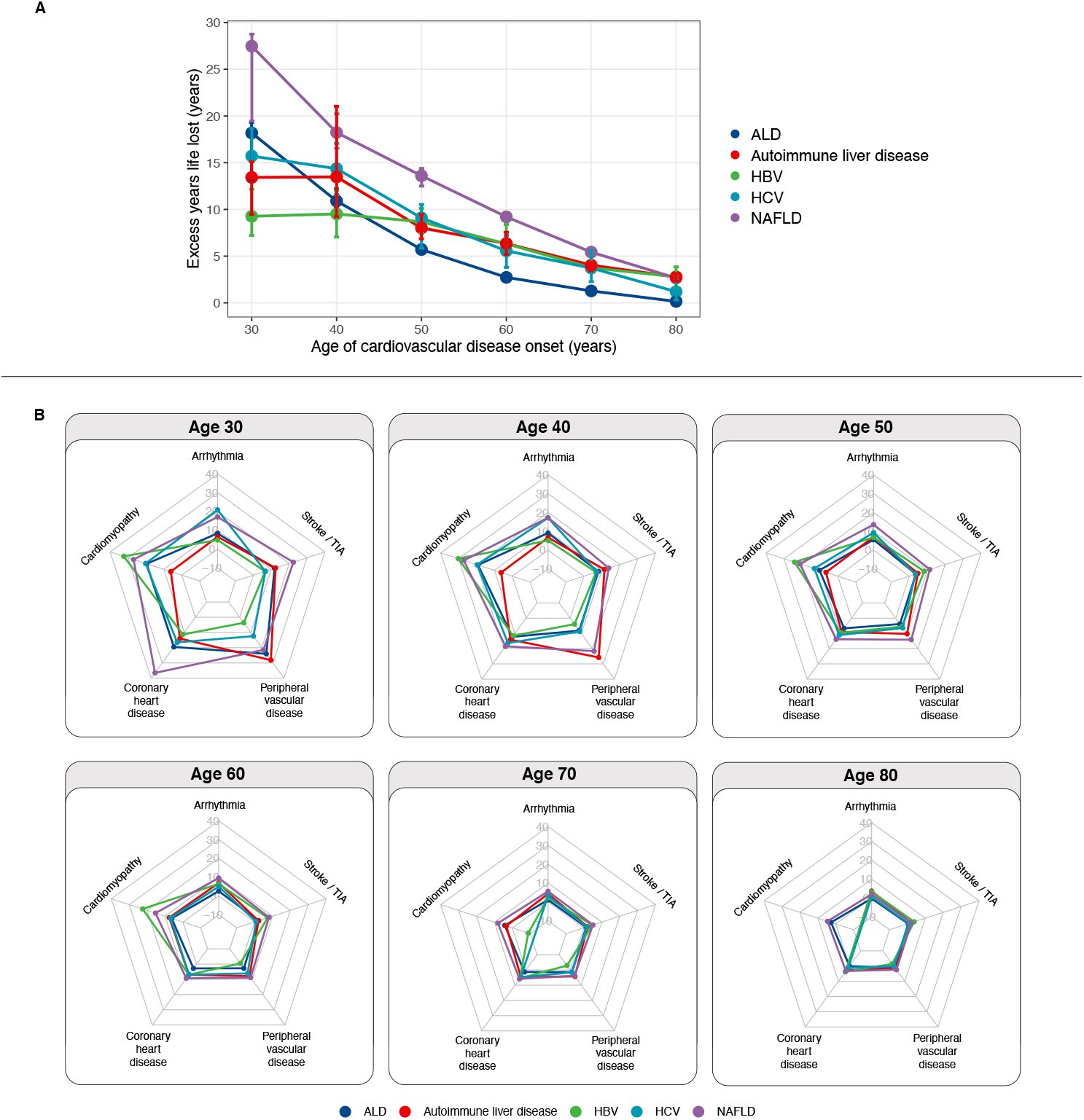
Excess years life lost (YLL) attributable to CVD in patients with liver disease. (A) Excess YLL for any initial presentation of CVD for different age of onset. (B) Excess YLL by each of the five CVD group for different age of onset.

When investigating sex differences, the difference in excess YLL between men and women appeared to be marginal in patients with ALD, autoimmune liver disease and NAFLD (Additional file 3). In patients with HBV, women diagnosed with CVD at ages 70 or 80 experience higher excess YLL than men. At age 70, women lost 7.4 years (CI: 3.2 – 9.0) while men lost 1.3 years (CI: −0.9 – 4.3). At age 80, women lost 6.1 years (CI: 3.5 – 8.7) and men lost 0.2 years (CI: −2.4 – 1.0) (Additional file 3). In patients with HCV, women diagnosed with CVD at ages 30 and 40 experienced higher excess YLL than men. For example, at age 40, women lost 18.6 years (CI: 15.9 – 22.9) while men lost 12.2 years (CI: 10.7 – 15.2) (Additional file 3).

We also evaluated excess YLL by age of CVD onset for each of the five liver diseases stratified by the five CVD categories. In the interest of maintaining conciseness and to aid comparison across liver diseases, we graphically displayed results as radar plots (Figure 5B). Each radar showed excess YLL estimates for a specific age of CVD onset, with each spoke representing excess YLL for one CVD category. A line is drawn to connect the excess YLL values for each CVD category where each liver disease is represented by different coloured lines. All categories of CVD were associated with shorter remaining life expectancies. When evaluating the surface areas covered by the lines, as expected, the radar plots demonstrated that younger age of CVD onset had higher excess YLL (as represented by a larger surface area connecting the CVD categories) that was consistent across the five liver conditions. Conversely, as the age of onset increased, excess YLL decreased. The radar plots also facilitate the visualisation of the differences by liver disease and CVD category. Notably, when comparing across the five liver conditions, the differences in excess YLL by CVD category were more pronounced at younger ages (Figure 5B). At age 40, patients with autoimmune liver disease, compared to other liver diseases, experience the highest excess YLL when diagnosed with peripheral vascular disease (25.7 years; CI: 12.7 – 41.6) compared to other CVD categories. In contrast, patients with autoimmune liver disease who are diagnosed with incident arrythmia at age 40 lost only 6.7 years (9.5% CI: 4.5 – 9.5) (Figure 5B).

We developed a public online tool to share results interactively (https://lailab.shinyapps.io/cvd_in_liver_disease/).

## Discussion

Our study examines incidence, comorbidity patterns, risk of initial presentation of CVD and excess YLL associated with CVD in patients with any of the five liver diseases. Patients with liver disease have an enhanced incidence burden of CVD compared to the general population. Coronary heart disease and arrhythmia are two of the most common first presentation of CVD, which show similar patterns across all liver diseases. This finding is consistent with previous studies in patients with NAFLD reporting an increase in the prevalence of coronary heart disease (including myocardial infarction), cardiac arrhythmias and cardiomyopathy^29^. Interestingly, the different aetiology of liver disease did not impact upon CVD presentation, perhaps suggesting that the liver disease drives CVD rather than the causative aetiology (i.e., viral infection or alcoholism). We provide the public, researchers and policymakers with an interactive online tool for exploration and visualisation of the incidence rates of liver disease, incidence rates of CVD in patients with and without liver disease and excess YLL associated with CVD. The tool also brings together information on other analyses such as comorbidity patterns in patients with liver disease and adjusted hazard ratios for CVD risk by age, biomarkers and comorbidities.

Our results highlight significant variations in the burden of liver disease across geographical regions in England. This suggests that there might be geographical variations in risk factors, health and risk awareness among the public, and access to diagnostic services where local solutions are required. Our observation that incidence rates for liver disease (except for HBV) were the highest in North East and North West regions was corroborated by the Public Health England’s (PHE’s) second atlas of variation for liver disease^30^. The PHE atlas revealed that Northern regions had a high rate of expenditure for hepatobiliary problems, liver disease admissions rates, liver disease mortality and rate of years of life lost. We observed that the incidence rate for HBV was the highest in London. This result was consistent with the PHE atlas which found that the percentage of women who tested positive for HBV during pregnancy screening and the rate of hospital admission for HBV-related end-stage liver disease and mortality were high in London, presumably caused by a high prevalence of migrant communities in this region^30^.

When examining the regional variations of CVD in patients with or without liver disease, we found that variation in incidence rates is ubiquitous across England. Our results draw attention to the need to coordinate CVD and liver disease services across regions to maintain equity in service access at a local level by helping commissioners, service providers and clinicians to compare the disease burden of their region with the national figure to ensure that care pathways are planned optimally. Regional variations signify that a one-size-fits-all approach is less likely to be effective. Services within each region need to identify challenges and adapt solutions at different rates in different scenarios. Our maps of disease burden variation could serve as a tool for benchmarking where each region stands relative to other regions and to provide a starting point for further investigation on reasons explaining the variation.

Our population-wide study of nearly 4 million individuals provides evidence that associations between clinical factors and 17 cardiovascular disease outcomes differ in terms of magnitude of effects when comparing across the five liver diseases. Raised BMI and current and former smoking were associated with an increased risk of CVD. Similarly, diabetes mellitus, hypertension, dyslipidaemia and chronic kidney disease are comorbidities associated with a higher risk. CVD risk is increased by at least two times in patients with impaired renal function^12^. We also observed that proteinuria, which is an indication of kidney damage, is associated with a higher risk of CVD. C-reactive protein is a marker of systemic inflammation, where it is found to be a predictor of mortality in patients with liver disease^31^. We found that patients with raised C-reactive protein have an increased risk of CVD, which corroborates previous findings that inflammation may serve as a pathogenic mechanism for triggering atherothrombotic CVD events^32^. Low albumin levels were associated with a higher risk. An inverse association between albumin and CVD may be explained by mechanisms related to the pathogenesis of CVD and inflammation associated with hypoalbuminaemia^33^.

We found that markers of liver function (i.e., bilirubin, aspartate transaminase and gamma-glutamyltransferase) do not appear to be associated with CVD risk. Evidence on the association between these measurements and CVD risk has been mixed^34^. When analysing the effects of alanine aminotransferase (ALT) levels, we observed an inverse relationship between ALT and CVD risk. A systematic review and meta-analysis of 29 cohort studies demonstrated that there was limited evidence for association between ALT and CVD events^35^. In contrast, stratified analysis on cause-specific CVD endpoints demonstrated that ALT was inversely associated with coronary heart disease but positively associated with stroke^35^. A study on three large prospective cohorts (West of Scotland Coronary Prevention Study, the Prospective Study of Pravastatin in the Elderly at Risk, and the Leiden 85-plus Study) with ages ranging from 45 to 85 years, also described an inverse association between ALT and cardiovascular outcomes, even after adjusting for confounders^36^. The authors found no evidence of any association between higher levels of ALT and increased risk of clinical outcomes when examining ALT levels >27U/L. In contrast, other studies reporting raised ALT levels with adverse outcomes were employing much higher ALT levels^37^ (defined as 2 times the upper limit of normal) than in our present study (we defined raised ALT as ≥ 35 U/L) and in the study by Ford et al^36^. High ALT in the normal range was also found to be negatively associated with death from ischaemic heart disease^38^. This suggests that there might be potential variations of CVD risk based on ALT levels that are closer to the population median. Furthermore, another report suggested a possible U-shaped association of ALT with vascular risk^39^. Findings from our study along with others suggest that the relationship between ALT and CVD risk is more complex than currently appreciated and may be caused by differences in underlying aetiology. It is also important to note that ALT levels are not useful indicators to risk-stratify the severity of liver disease^40^. Patients with cirrhosis can have normal values when scar tissue replaces the damaged liver cells and can no longer produce ALT.

Our results confirm that patients with liver disease experience premature mortality attributable to CVD. These findings are consistent with a report from the Centers for Disease Control and Prevention stating that CVD ranks third in YLL prior to age 65^41^ along with other studies demonstrating that premature death due to CVD continues to increase globally^42,43^. Although each type of CVD is associated with excess YLL, we observed that women lost more years of life compared to men at a younger age of CVD onset among patients with HCV and at an older age of CVD onset among patients with HBV.

### Strengths and weaknesses

A major strength of this study is the ability to differentiate 17 cardiovascular outcomes using a real-world population cohort, which is larger, contemporary and more representative of the general population compared with investigator-led cohorts. Given that we have analysed records from both primary and secondary care, we are able to capture conditions that are treated by general practitioners and specialist clinicians in hospitals. Primary care records also provide more detailed data than those recorded on admission to hospital, where the former account for the total population while the latter only for a subset of individuals who attended hospitals. In addition, given the longevity of follow-up and the breadth and depth of variables available, we were able to examine the associations between a wide range of comorbidities and biomarkers. Another strength is our use of openly accessible codelists for reuse that are made available publicly (https://www.caliberresearch.org/portal) and previously validated in numerous studies^18,19,21^.

We note important limitations in this study. First, our study employs only data in England, which may limit generalisability to other geographical locations. We have only sampled a subset of English general practices accounting for almost 4 million individuals. Second, as in all observational studies, there is the potential for residual confounding. Third, missing data is a common phenomenon in EHRs, however, sensitivity analyses on complete records have shown that our estimates were robust to our assumptions around missing data.

### Policy implications

Our study has policy implications in CVD prevention and targeted recommendations. CVD prevention policies and guidelines should identify groups of high-risk individuals among people with liver disease for targeted screening and intervention. Treatment of HCV with direct-acting antiviral therapy is linked to a decrease in the risk of CVD events in patients who have sustained virological responses^44^. This suggests that HCV infection and potentially HBV infection may be modifiable CVD risk factors^45^ and treating the underlying infection could improve both liver and CVD outcomes.

This work has important implications for addressing inequalities in HBV and HCV screening and treatment. According to recent estimates, around 113,000 and 180,000 individuals are infected with HCV and HBV in England respectively^46^. Although effective treatment for HCV exists, there remains inequality in accessing treatment as HCV may remain undiagnosed in underserved populations such as people who are homeless, in prison or drug users. Migration from HBV endemic countries has resulted in an increased prevalence of HBV infection in the UK. However, migrants remained an underserved or invisible population due to low disease awareness and low engagement with health services^47^. Our results suggest that regional initiatives targeting underserved communities might help complement interventions from two perspectives – treatment of chronic HBV/HCV by addressing social challenges that cause healthcare inequality and targeted prevention of CVD in patients with liver disease.

### Conclusion

We propose that management of liver disease should include regular cardiovascular risk assessment in addition to checking for undiagnosed diabetes and renal comorbidities. Furthermore, CVD risk assessment should be provided for all types and all stages of liver disease. At the individual level, communicating risk information may affect people’s attitude to seek opportunities for early detection and treatment of CVD to reduce premature mortality. For example, this can be accomplished by improving access to the NHS Health Check programme^48^, aimed at individuals aged 40 to 74, to identify early signs of CVD, stroke, kidney disease and diabetes. Access to risk information could help stimulate conversations between patients and their doctors on decisions on managing their health and care. It is now possible to generate and disseminate novel risk information using real-world data that takes a step towards making decisions that are not based on the group average but that are closer to specific patient scenarios (i.e., patients like me) encountered in clinical settings.

## Data Availability

All data produced in the present work are contained in the manuscript.

## Availability of data and materials

The data used in this study are available on successful ethics application to the Clinical Practice Research Datalink (CPRD). All summarised data and results are made available as supplementary materials.

### Abbreviations

CVD: Cardiovascular disease
EHR: Electronic health records
ALD: Alcoholic liver disease
HBV: Chronic hepatitis B infection
HCV: Chronic hepatitis C infection
NAFLD: Non-alcoholic fatty liver disease
YLL: Years of life lost
INR: International Normalised Ratio
TIA: Transient ischaemic attack
HR: Hazard ratio
CI: Confidence interval
ALT: Alanine aminotransferase

## Contributions

Research question: WHC and AGL

Funding: AGL

Study design and analysis plan: WHC, SM and AGL

Preparation of data: WHC, SM and AGL

Statistical analysis: WHC, SM and AGL

Creation of online app: SM

Drafting initial and final versions of manuscript: WHC and AGL

Critical review of early and final versions of manuscript: All authors

## Ethics declarations

### Ethics approval and consent to participate

Information governance approval was obtained from the Medicines Healthcare Regulatory Authority (UK) Independent Scientific Advisory Committee (21_000363) Clinical Practice Research Datalink (CPRD).

### Consent for publication

Not applicable

### Competing interests

GRF receives funding from companies that manufacture drugs for hepatitis C virus (AbbVie, Gilead, MSD) and consults for GSK, Arbutus and Shionogi in areas unrelated to this research.

## Acknowledgements

Not applicable.

## Funding

AGL is supported by funding from the Wellcome Trust (204841/Z/16/Z), National Institute for Health Research (NIHR) University College London Hospitals Biomedical Research Centre (BRC714/HI/RW/101440), the Academy of Medical Sciences (SBF006\1084), NIHR Great Ormond Street Hospital Biomedical Research Centre (19RX02) and the Health Data Research UK Better Care Catalyst Award (CFC0125). The funders had no role in study design, data collection and analysis, decision to publish, or preparation of the manuscript.

## Supplementary results

### Regional variations in age-standardised and age-specific incidence rates for liver disease

An exploration of regional variations revealed that the patterns of incidence burden differed across liver diseases (Additional file 5). North West England had the highest age-standardised incidence rate for any liver disease (151 per 100,000 person years), while the lowest incidence rate was observed in Yorkshire and the Humber (87 per 100,000 person years) (Figure 1A, Additional file 5).

Delving into specific liver diseases, regions with the highest incidence rates were as follow: ALD (North West; 35 per 100,000 person years), autoimmune liver disease (North East; 6 per 100,000 person years), HBV (London; 11 per 100,000 person years), HCV (North East; 10 per 100,000 person years) and NAFLD (London; 116 per 100,000 person years). The North West and North East regions consistently exhibited relatively high age-standardised incidence rates for individual liver diseases (except for HBV), while the regions in the east of England (i.e., Yorkshire and The Humber, East Midlands and East of England), had consistently low incidence rates (Figure 1A, Additional file 5). For HBV, however, we found that London had a much higher incidence rate compared to all other regions (11 in London vs. 0.8 to 3.6 per 100,000 person years in other regions).

Regionally, the maximum incidence for any liver disease was reached in individuals aged 50-59 for all regions except the West Midlands (where incidence peaked in individuals age 60-69) (Additional file 6). Additional age-specific incidence rates by specific liver disease type and geographical regions were provided in Additional file 6.

### Regional variations in age-standardised and age-specific incidence rates for CVD in patients with or without liver disease

When exploring regional differences in patients with liver disease, individuals from the North East (3,179 per 100,000 person years, CI: 2,423-3,935) and North West (2,960 per 100,000 person years, CI: 2,704-3,217) had the highest and second highest incidence burdens for CVD, respectively (Figure 2A, Additional file 7). Similarly, the North West region had the highest incidence for CVD in people without liver disease (1,537 per 100,000 person years) (Figure 2B, Additional file 8). In individuals with liver disease, the lowest CVD incidence was observed in the South East region (2,420 per 100,000 person years, CI: 2,098-2,742) (Figure 2A, Additional file 7). However, in people without liver disease, East of England had the lowest CVD incidence rate (1,226 per 100,000 person years) (Figure 2B, Additional file 8).

Variation rates were more striking when comparing liver disease types. Regions with the highest incidence burdens of CVD by liver disease type were as follow: ALD (North East; 3,827 per 100,000 person years), autoimmune (North East; 3,512 per 100,000 person years), HBV (West Midlands; 3,119 per 100,000 person years), HCV (South Central; 3,714 per 100,000 person years) and NAFLD (East Midlands; 3,325 per 100,000 person years) (Figure 2A, Additional file 7).

When investigating regional variations in age-specific incidence rates, the highest CVD incidence in people with and without liver disease was observed in individuals aged 70 and above across all regions (Additional file 9, Additional file 10).

### Comorbidity patterns in patients with liver disease

We analysed comorbidity patterns for 14 prevalent conditions stratified by liver disease type, age group and sex (Additional file 1). Comorbidities were common in people with liver disease. A general trend of increased in diagnosed comorbidities was observed across all liver diseases. Men with autoimmune liver disease had an increased prevalence of Crohn’s disease especially in the younger age groups: age 30-39 (8.3%) and age 40-49 (11.8%) (Additional file 1). The prevalence of Barrett’s oesophagus, renal disease, diabetes mellitus, complications of diabetes, diverticular disease of the intestine, dyslipidaemia and hypertension exhibited an upward trend with increasing age and displayed consistent patterns across the five liver conditions. Among patients with HBV at ages 70-79, diabetes was present in 31.5% men and 29.0% women, whereas, in patients aged 40-49, the proportions were 7.2% and 6.5% in men and women respectively. Similarly, among patients with NAFLD, renal disease was present in 18.3% men and 23.5% women of ages 70-79, whereas, in patients aged 40-49, only 3.3% men and 8.6% women had renal disease (Additional file 1).

## Notes

### Author Declarations

Ethics approval was obtained from the Medicines Healthcare Regulatory Authority (UK) Independent Scientific Advisory Committee (21_000363) Clinical Practice Research Datalink (CPRD).

